# Epidemiology of pathologic myopia in UK adults with high myopia

**DOI:** 10.1101/2024.11.19.24317505

**Authors:** Fabian Yii, Niall Strang, Miguel O. Bernabeu, Baljean Dhillon, Tom MacGillivray, Ian J.C. MacCormick

## Abstract

**Aims:** To conduct the first cross-sectional epidemiological investigation of pathologic myopia (PM) in UK adults with high myopia.

**Methods:** Fundus photographs of 3,024 highly myopic eyes (spherical equivalent refraction, SER ≤ –5.00D) from 2,000 randomly sampled adults (aged 40-70 years) in the UK Biobank were double graded by an ophthalmic reading centre using the Meta-analysis for Pathologic Myopia framework. Adjudication was performed by one of two retinal specialists. Multivariable mixed-effects logistic regression was used to explore potential risk factors and fundus biomarkers—initially controlling for SER, age and sex, before including all variables with p<0.10 in a single model.

**Results:** PM was present in 1,138 of 3,006 gradable fundus photographs, with 41.7% (95% CI: 39.5%-43.9%) of participants affected in at least one eye graded. Most eyes with PM exhibited diffuse chorioretinal atrophy (97.4%), while the more severe stages— patchy chorioretinal atrophy and macular atrophy—were observed in only 24 and 5 eyes, respectively. Thirteen eyes had “plus” lesions or suspected staphyloma. Factors independently associated with increased odds of PM (all p<0.05) included decreasing SER (adjusted odds ratio: 0.22, 95% CI: 0.15-0.32), older age (2.20, 1.63-2.97), female sex (1.87, 1.12-3.12), lower deprivation (0.73, 0.56-0.94), White ethnicity (52.3, 17.3-158.3), lower retinal arteriovenous ratio (0.47, 0.37-0.58), increased retinal vascular complexity (4.68, 3.22-6.81) and a relatively horizontal disc orientation (2.98, 1.88-4.72). None of the explored modifiable lifestyle or health-related variables were associated with PM.

**Conclusions:** PM prevalence is high among mid-life adults with high myopia in the UK Biobank. Although most cases are mild, the progressive and age-related nature of PM means an elevated risk of irreversible visual impairment for these individuals in later life.

## Introduction

While myopia has traditionally been regarded as a relatively benign vision disorder requiring little more than optical correction, there is growing recognition of its insidious and far-reaching implications for ocular health.^1^ One important myopic sequela is pathologic myopia (PM), which has in recent years been identified as the leading cause of irreversible blindness among adults in parts of Asia.^2^ In the United Kingdom (UK), its impact was noted as early as seven decades ago in a report by Sorsby (cited in Duke-Elder’s celebrated *System of Ophthalmology*)—where after reviewing 36,617 certificates of blindness issued between 1951 and 1954, “pathological myopia” was identified as the commonest cause of legal blindness in English adults aged 40-60 years.^3, 4^

Due to its largely irreversible and visually debilitating consequences, the global economic burden of PM is substantial, reaching approximately 6 billion USD in 2015 based on conservative estimates.^5^ The estimated prevalence of PM in population-based studies (almost all conducted in Asia) ranges from 25.3% to 71.4% in highly myopic adults (spherical equivalent refraction, SER ≤ –5D or –6D), with a meta-analysed prevalence of 47.4%.^6, 7^ However, there is a paucity of data on the epidemiology of PM in the UK. Knowledge about its risk factors and imaging biomarkers also remains limited, hindering personalised risk stratification in clinical practice.^8^ The UK Biobank (ukbiobank.ac.uk) is a large-scale, population-based cohort of middle-aged adults recruited from 22 sites across the UK. With its extensive breadth and depth, the dataset provides a unique opportunity to capture a detailed snapshot of PM at the population level. Using this dataset, we aimed to conduct the first epidemiological investigation of PM in the UK, determining its prevalence and risk factors among adults with high myopia (defined as SER ≤ –5D).

## Methods

### Cohort description

Study participants were derived from the UK Biobank Eye and Vision sub-cohort, which comprised 68,508 participants who underwent an extensive range of assessments, including various ophthalmic tests between 2009 and 2010. Detailed description of the dataset is available elsewhere.^9^ Briefly, the ophthalmic assessments included 45° macula-centred colour fundus photography using the Topcon 3D OCT1000 Mark II (Topcon Corporation, Tokyo, Japan), autorefracto-keratometry without cycloplegia using the Tomey RC-5000 (Tomey, Nagoya, Japan), distance visual acuity (VA) assessment with habitual distance correction (if required) using the Precision Vision digital LogMAR chart (Precision Vision, LaSalle, USA) and intraocular pressure (IOP) measurement using the Ocular Response Analyzer (Reichert Corporation, Philadelphia, USA).

Figure 1 provides an overview of the sample selection process applied in this study. After removing eyes without fundus photographs and those with “reject” image quality, as determined using a validated deep learning model,^10^ 90,191 eyes of 51,534 participants remained. We further excluded eyes with missing refractive error, leaving 89,216 eyes of 51,086 participants. Among these, 3,821 participants had high myopia in at least one eye (SER ≤ –5.00D). We randomly sampled 2,000 of these participants and graded the fundus photographs of 3,024 eyes meeting the definition of high myopia outlined above.

**Figure 1.**
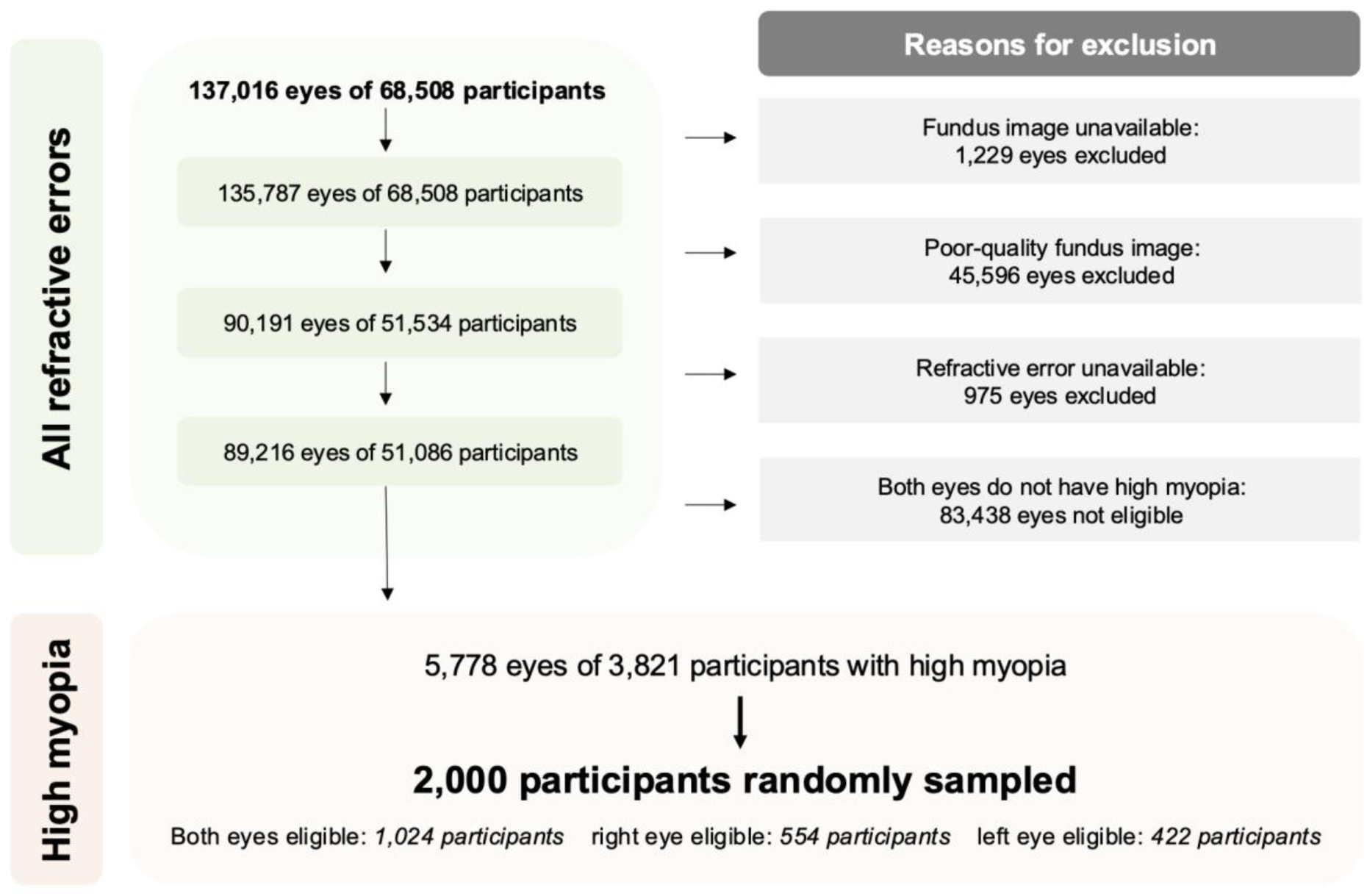
Flowchart providing an overview of the sample selection process, in which 2,000 participants were randomly sampled from a population of 3,821 participants with high myopia (spherical equivalent refraction < –5.00D) in at least one eye. Among those sampled, both eyes of 1,204 participants, the right eye of 554 participants and the left eye of 422 participants had high myopia, making these eyes eligible for grading.

### Pathologic myopia grading

Each fundus photograph was independently graded by a pair of ophthalmic graders at the Belfast Ophthalmic Reading Centre (networcuk.com/Home/Belfast), using the Meta-analysis for Pathologic Myopia classification framework (META-PM).^11^ Myopic maculopathy, a key clinical manifestation of PM, is categorised according to the framework as follows: no myopic maculopathy (M0), fundus tessellation only (M1), diffuse chorioretinal atrophy (M2), patchy chorioretinal atrophy (M3) and macular atrophy (M4). M2 can be further divided into non-macular and macular subtypes, with the former generally preceding the latter.^2, 12^ Myopic maculopathy is defined as M2 or higher. Examples of images for each category of myopic maculopathy graded in the present study are available as Supplementary Figure 1.

Three “plus” lesions (lacquer cracks, myopic choroidal neovascularisation and Fuchs’ spot), which can coexist with any category of myopic maculopathy, are also part of the META-PM definition. We defined PM as the presence of any of the following: myopic maculopathy, “plus” lesion(s) or suspected posterior staphyloma. One of two consultant retinal specialists adjudicated each instance of inter-grader disagreement. All graders, including the adjudicators, were masked to participant characteristics and had access only to the fundus photographs throughout the entire process.

### Risk factors

Unless otherwise stated, the following demographic, socioeconomic or lifestyle variables were based on a self-administered touchscreen questionnaire completed on the day of assessment. The Townsend deprivation index, a continuous (numeric) variable, measured relative deprivation in a given postcode area using national census data, where a more positive value indicated a greater degree of deprivation.^13^ Education level was binarised into post-secondary education and secondary/lower education. Ethnicity was also binarised—into “non-White” and “White”—as the cohort was predominantly (>90%) White British or from other White backgrounds. Lifestyle variables included smoking status (binarised into never/previous and current), alcohol consumption (binarised into frequent and infrequent, with “daily or almost daily” as threshold) and sleep duration (number of hours per day as an integer).

Hypertension was defined using linked healthcare data^14^ or if either the systolic or diastolic blood pressure, measured with the Omron HEM-705IT (Omron Corporation, Kyoto, Japan), was equal to or exceeded 140mmHg or 90mmHg, respectively, on the day of assessment. Cardiovascular disease was defined based on linked healthcare data^14^ as a history of myocardial infarction, cardiomyopathy, ischaemic heart disease, cardiac arrest, heart failure, multiple valvular heart disease, atherosclerosis or any other heart condition. Diabetes was also defined based on linked healthcare data^14^ or if random glucose, measured by hexokinase analysis on the Beckman Coulter AU5800 (Beckman Coulter, Brea, USA), exceeded 11.1mmol/L (equivalent to 200 mg/dl) on the day of assessment. Body mass index (BMI, in Kg/m^2^) and total cholesterol (in mmol/L) were derived from anthropometric measurements using standard scales and CHO-POD analysis on the Beckman Coulter AU5800 (Beckman Coulter, Brea, USA), respectively. Both variables were analysed as continuous variables. Explored ocular variables included glaucoma status, derived from linked healthcare data,^14^ and IOP.

### Fundus biomarkers

Several imaging features were derived from fundus photographs using methods detailed previously,^15^ focussing on dimensionless metrics to avoid issues related to camera telecentricity and ocular magnification.^16^ Vascular metrics included fractal dimension (FD), tortuosity, temporal arterial/venous concavity and arteriovenous ratio (AVR), all of which were expressed as continuous variables. A greater FD value suggested a more complex retinal vasculature, while a larger tortuosity value indicated increased tortuosity. Temporal arterial/venous concavity described the overall parabolic course of the vessels, with a larger value indicating a more inward course of the major temporal artery/vein towards the fovea. AVR was calculated as the ratio of central retinal arteriolar equivalent to central retinal venous equivalent.

The morphology of the optic disc (OD) was captured by measurements of its tilt (also known as ovality) and orientation. Tilt was the ratio of major axis length to the minor axis length, while orientation was the absolute angle between the horizontal axis of the image and the major axis of the OD. Both metrics were binarised for interpretability: a tilt value ≥1.3 suggested the OD was tilted,^17^ while an orientation angle ≥45° meant the OD was relatively vertically orientated. Recognising the challenges in accurately localising the fovea in cases of moderate or severe PM, no metrics requiring foveal localisation were derived.

### Statistical analysis

Multivariable logistic regression was used to explore potential risk factors and fundus biomarkers (as introduced above) by initially fitting separate models with each variable as the sole independent variable, while controlling for SER, age and sex. All variables with p<0.10 were subsequently included in a single (final) model—while similarly controlling for SER, age and sex—to assess their independent associations with PM. All models included random intercepts with individuals as random effects to account for inter-eye correlation, fitted using the *glmer* function available from the *lme4* package in R version 4.2.2 (R Core Team 2022, Vienna, Austria). Continuous variables were all standardised to have zero mean and unit variance during model fitting to ensure model stability, as some variables were on very different scales. Multicollinearity was checked using variance inflation factor, treating 10 as the cutoff value.^18^ The source code is freely and openly available at github.com/fyii200/UKbiobankPM.

## Results

### Participant characteristics

Eighteen of the 3,024 assessed fundus photographs were ungradable due to image quality issues: 7 were unanimously rejected by both graders at the reading centre and 11 were further rejected during adjudication (examples and reasons are given in Supplementary Figure 2), leaving 3006 eyes of 1994 participants for analysis. Table 1 presents the summary statistics of key variables for the gradable and ungradable eyes.

**Table 1.** Summary statistics of key variables (mean ± standard deviation shown for continuous variables). Eye-specific variables are summarised at the eye level, while participant-specific variables are summarised at the individual level.

| Variable | Gradable |  |  | Ungradable<br>(18 eyes) |
| --- | --- | --- | --- | --- |
|  | Overall<br>(3,006 eyes) | Normal<br>(1,868 eyes) | Pathologic<br>(1,138 eyes) |  |
| Spherical equivalent refraction (D) | -7.31 ± 2.39 | -6.90 ± 1.82 | -7.99 ± 2.97 | -8.34 ± 4.2 |
| Age (year) | 54.7 ± 7.7 | 54.5 ± 7.8 | 56.0 ± 7.5 | 60.5 ± 5.9 |
| Female | 58.7% | 58.1% | 59.9% | 72.2% |
| Townsend deprivation index | -0.97 ± 2.90 | -0.84 ± 2.94 | -1.20 ± 2.85 | -0.76 ± 3.30 |
| > Secondary education | 60.0% | 60.4% | 60.2% | 50.0% |
| White ethnicity | 91.1% | 87.6% | 97.7% | 100% |
| Never or previously smoked | 93.4% | 92.4% | 95.1% | 88.9% |
| Alcohol: daily or almost daily | 19.6% | 19.9% | 19.7% | 16.7% |
| Sleep duration (hours per day) | 7.1 ± 1.0 | 7.1 ± 1.0 | 7.2 ± 1.0 | 6.9 ± 1.1 |
| Hypertension | 20.4% | 20.3% | 20.3% | 28.6% |
| Cardiovascular disease | 3.2% | 2.8% | 3.6% | 0% |
| Diabetes | 4.0% | 4.6% | 3.2% | 5.6% |
| Body mass index (kg/m<sup>2</sup>) | 26.8 ± 4.8 | 26.7 ± 4.7 | 27.0 ± 5.0 | 25.6 ± 4.1 |
| Total cholesterol (mmol/L) | 5.7 ± 1.1 | 5.7 ± 1.1 | 5.8 ± 1.1 | 6.2 ± 1.0 |
| Glaucoma | 2.4% | 2.4% | 2.4% | 0% |
| Intraocular pressure (mmHg) | 16.5 ± 3.7 | 16.3 ± 3.6 | 16.7 ± 3.9 | 16.0 ± 3.4 |
| Vessel fractal dimension | 1.44 ± 0.07 | 1.44 ± 0.07 | 1.45 ± 0.06 | NA |
| Vessel tortuosity | 0.69 ± 0.04 | 0.69 ± 0.04 | 0.69 ± 0.04 | NA |
| Temporal arterial concavity | 0.003 ± 0.001 | 0.003 ± 0.001 | 0.003 ± 0.001 | NA |
| Temporal venous concavity | 0.003 ± 0.001 | 0.003 ± 0.001 | 0.003 ± 0.001 | NA |
| Retinal arteriovenous ratio | 0.65 ± 0.08 | 0.66 ± 0.08 | 0.63 ± 0.08 | NA |
| Tilted optic disc | 2.4% | 1.7% | 3.6% | NA |
| Horizontally orientated optic disc | 26.1% | 21.5% | 33.6% | NA |

In general, participants with ungradable images tended be older females with higher myopia. White ethnicity accounted for 1,817 (91.1%) of the 1994 participants included in the analysis. Most of the remaining 177 non-White participants were from Indian/Pakistani (N=56), Chinese (N=43) and Caribbean (N=42) backgrounds. The inter-grader agreement was high, with a weighted kappa coefficient of 0.80 and an overall agreement of 84%. Most adjudications were attributable to disagreements between M1 and non-macular M2 (50%), followed by non-macular M2 and macular M2 (28%), and M0 and M1 (9%).

### Disease prevalence

Overall, 1,138 out of 3,006 gradable fundus photographs showed signs of PM (37.9%, 95% CI: 36.1% to 39.6%), with 41.7% (95% CI: 39.5% to 43.9%) of participants affected in at least one eye graded. The majority (1,107 eyes) exhibited the earliest stage of myopic maculopathy (M2), with the non-macular subtype being more prevalent than the macular subtype (648 eyes vs 459 eyes). Both patchy chorioretinal atrophy and macular atrophy were rare in this dataset, affecting only 24 (0.8%) and 5 (0.2%) eyes, respectively. Among eyes without myopic maculopathy, 1,780 (59.2%) exhibited varying degrees of fundus tessellation.

Twelve eyes had “plus” lesions, with the majority of eyes (10) showing concurrent myopic maculopathy. Fuchs’ spot was found in 2 eyes M1, 1 eye with non-macular M2 and 1 eye with M3. Lacquer cracks were present in 5 eyes with macular M2, 1 eye with non-macular M2 and 1 eye with M3. Only 1 eye was suspected to have posterior staphyloma, which coexisted with patchy chorioretinal atrophy. The median (interquartile range) VA for eyes with M0, M1, non-macular M2, macular M2 and M3 were 0 (0.18), 0 (0.18), 0.02 (0.2), 0.06 (0.22) and 0.24 (0.5) logMAR, respectively. Note that VA was available for only 1 (0.34 logMAR) out of the 5 eyes with M4 due to challenges in assessing VA in these eyes lacking good central fixation.

### Risk factors and fundus biomarkers

Figures 2 and 3 show the percentages of eyes with PM—stratified by demographic /socioeconomic variables and fundus features associated with PM in the final multivariable model (Table 2). The variance inflation factor was low (≤ 2.2) for all variables in the final model, suggesting no major issues with multilinearity. As expected, higher myopia and older age were independently associated with increased odds of PM (Table 2). To illustrate, PM prevalence was lowest (25.4%, 95% CI: 20.6% to 30.7%) in eyes with myopia below –6.00D among younger participants aged 40-50 years, whereas the highest prevalence (67.9%, 95% CI: 58.4% to 76.4%) was observed in eyes with myopia ≤ –10.00D among older participants aged 60-70 years (Figure 2). Females had a higher overall prevalence of PM (42.5%, 95% CI: 39.7% to 45.4%) than males (40.5%, 95% CI: 37.1% to 43.9%), although this only appeared evident at higher levels of myopia (Figure 2) and in the final model adjusting for other covariates (Table 2).

**Figure 2.**
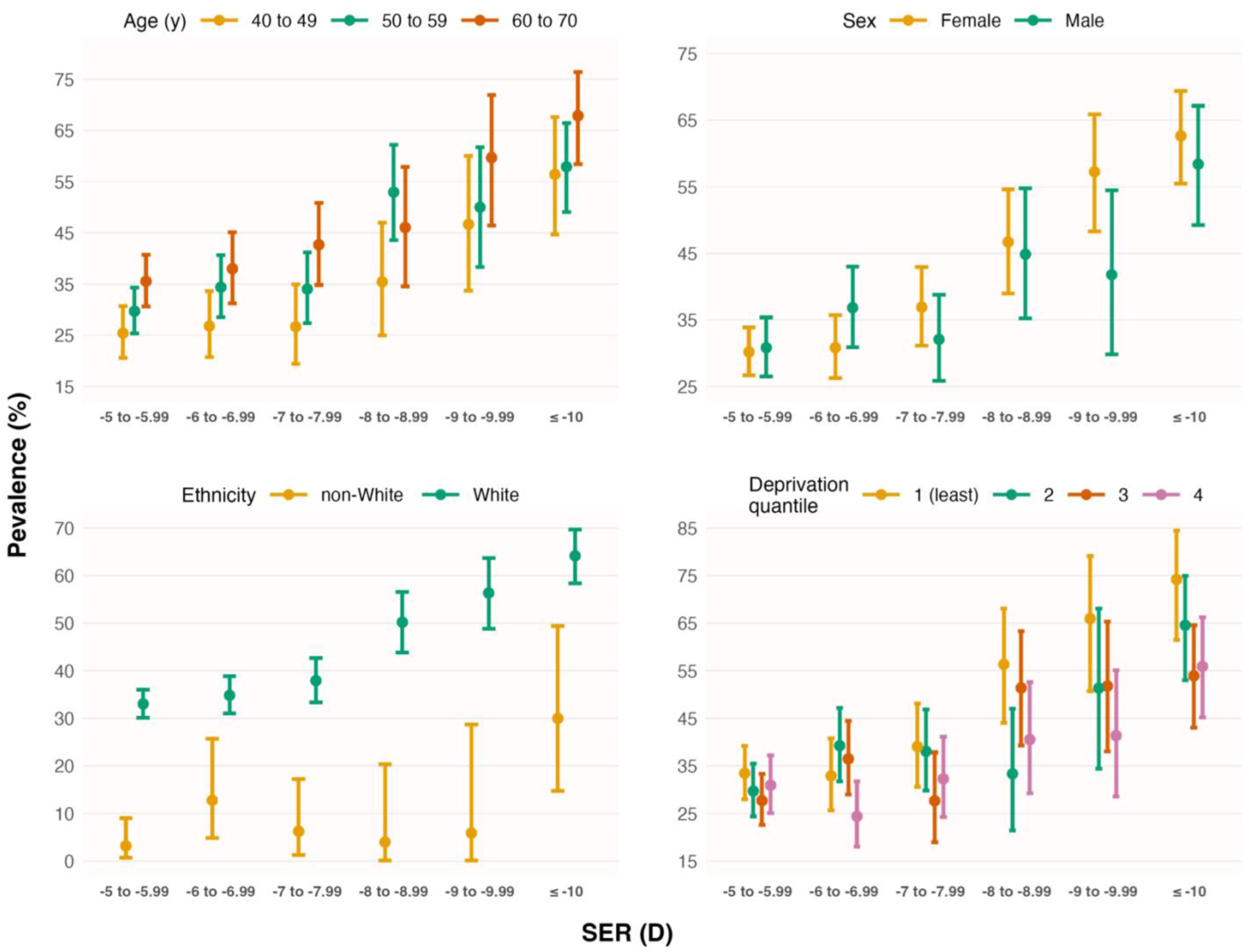
Prevalence (± 95% confidence intervals) of pathologic myopia vs spherical equivalent refraction (SER), stratified by age, sex, ethnicity and Townsend deprivation index (divided into 4 quantiles for illustration purposes).

**Figure 3.**
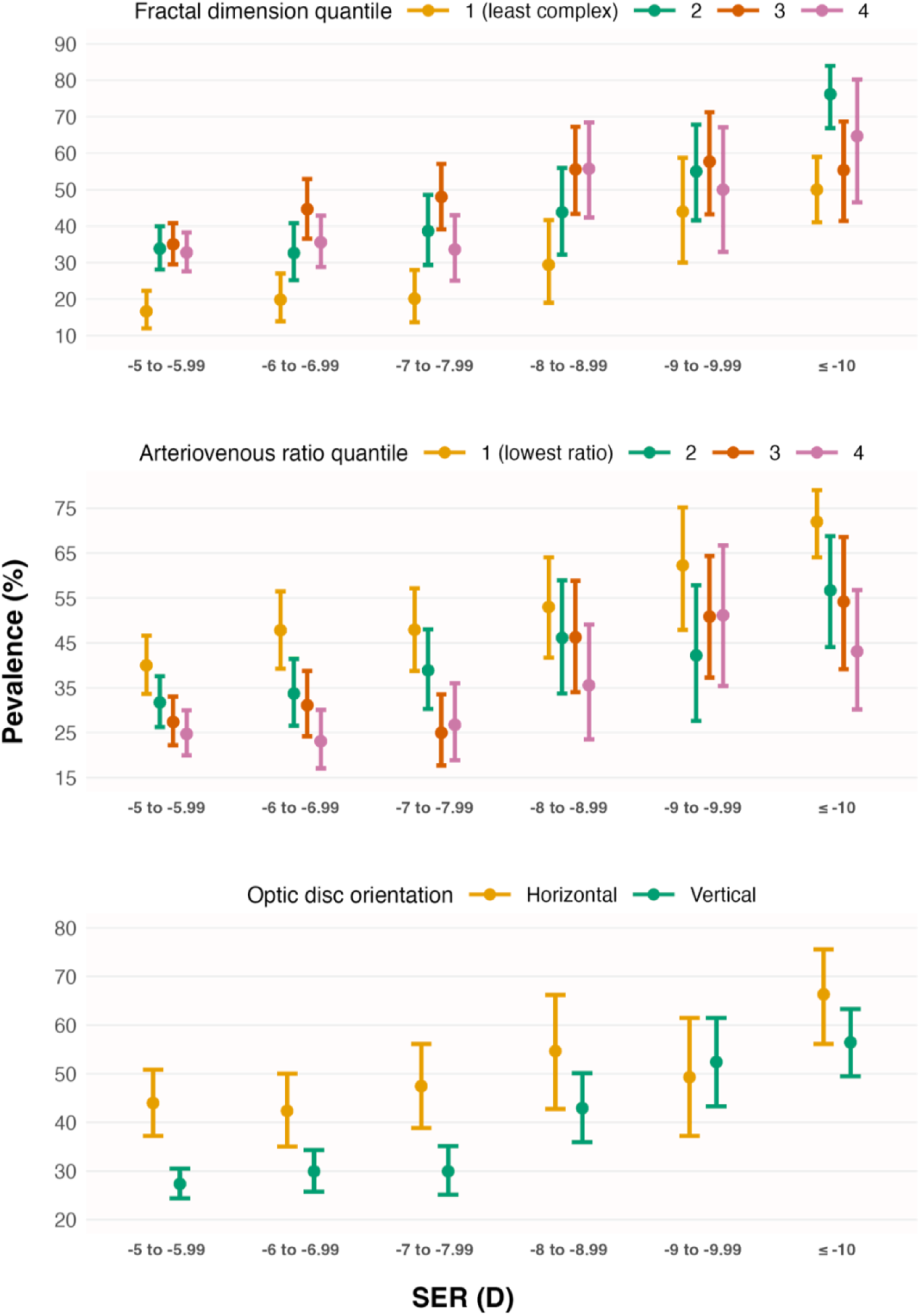
Prevalence (± 95% confidence intervals) of pathologic myopia vs spherical equivalent refraction (SER), stratified by vessel fractal dimension (divided into 4 quantiles for illustration purposes), retinal arteriovenous ratio (divided into 4 quantiles for illustration purposes) and optic disc orientation (angle between horizontal axis of the image and major axis of the disc).

**Table 2.** Multivariable logistic regression with pathologic myopia (binary) as the dependent variable, initially controlling for spherical equivalent refraction (SER), age and sex (left), before including all variables with p<0.10 in a single model (right). All continuous variables are standardised to have zero mean and unit variance. Regression results for non-significant variables are shown in Supplementary Table 1.

| Variable | Adjusted for SER, age and sex |  | Final model |  |
| --- | --- | --- | --- | --- |
|  | Odds ratio [95% CI] | P | Odds ratio [95% CI] | P |
| SER | 0.22 [0.12 to 0.40] | <0.001 | 0.22 [0.15 to 0.32] | <0.001 |
| Age | 1.74 [1.21 to 2.51] | <0.01 | 2.20 [1.63 to 2.97] | <0.001 |
| Female | 1.30 [0.64 to 2.65] | 0.47 | 1.87 [1.12 to 3.12] | 0.02 |
| Townsend deprivation index | 0.71 [0.49 to 1.02] | 0.06 | 0.73 [0.56 to 0.94] | 0.01 |
| White ethnicity | 43.1 [17.0 to 109.7] | <0.001 | 52.3 [17.3 to 158.3] | <0.001 |
| Vessel fractal dimension | 6.44 [4.20 to 9.87] | <0.001 | 4.60 [3.16 to 6.70] | <0.001 |
| Retinal arteriovenous ratio | 0.59 [0.44 to 0.79] | <0.001 | 0.47 [0.37 to 0.58] | <0.001 |
| Horizontally orientated disc | 2.65 [1.50 to 4.69] | <0.001 | 2.98 [1.88 to 4.72] | <0.001 |

Additionally, White ethnicity was associated with increased odds of PM—regardless of myopia severity and other potential socioeconomic confounders (Figure 2 & Table 2). Within the non-White ethnic group, PM prevalence was 7.7% (95% CI: 1.6% to 20.9%) among Indian/Pakistani participants, 3.3% (95% CI: 0.1% to 17.2%) among Chinese participants and 3.7% (95% CI: 0.1% to 19.0%) among Caribbean participants—lower than the 44.7% (95% CI: 42.4% to 47.0%) observed in White participants. Participants from less deprived areas, as indicated by a more negative Townsend deprivation index, also had higher odds of PM (Figure 2 & Table 2). Fundus features found to be independently associated with increased odds of PM included greater vessel FD, lower AVR and a relatively horizontally orientated OD (Table 2). While these imaging features were associated with PM irrespective of myopia severity, the effects appeared strongest in eyes with myopia between –5.00D and –8.00D (Figure 3). Regression results for other non-significant variables—such as education level, smoking status, alcohol consumption and health-related variables—are presented in Supplementary Table 1.

## Discussion

The prevalence of PM among highly myopic adults in the UK Biobank is high, with slightly over 40% affected in at least one eye graded. Most (97.4%) eyes with PM exhibited the earliest stage—diffuse chorioretinal atrophy—with the milder non-macular subtype (58.5%) being more prevalent than the macular subtype. Patchy chorioretinal atrophy and macular atrophy affected only 1% of all eyes with high myopia. Higher myopia, older age, female sex, White ethnicity, lower deprivation, higher vessel complexity, lower AVR and a relatively horizontal OD orientation were all independently associated with increased odds of PM. None of the modifiable lifestyle or health-related variables explored herein were associated with PM.

Among the few epidemiological studies conducted on populations of European ancestry, PM prevalence was also found to be high among highly myopic (≤ –6D) Dutch adults: 25.0% 24.5% and 35.2% among those aged 40-49, 50-59 and 60-69 years, respectively.^19^ In a German sample of 519 high myopes from the Gutenberg Health Study, aged 35-74 years, the prevalence of PM was reported to be lower, around 10%.^20^ Despite variations in prevalence, these studies similarly noted that most cases were relatively mild, with diffuse chorioretinal atrophy being the commonest presentation. In an older population-based study examining the prevalence of myopic retinopathy—defined as the presence of staphyloma, lacquer cracks, Fuchs’ spot or myopic chorioretinal atrophy—in predominantly White Australians aged ≥ 49 years, the prevalence was 25% for SER ≤ –5D, with more than 50% of eyes with myopia above – 9D affected.^21^ Likewise, among adults aged ≥ 40 years in the Beijing Eye Study, the prevalence of myopic retinopathy was found to be very high in 214 highly myopic eyes (≤ 6D): 40.4% for myopia between –6D and –7.99D and 89.6% for myopia ≤ –10D.^22^ In the Singapore Epidemiology of Eye Diseases (SEED) cohort, PM was reported to affect 28.7% of 523 highly myopic (≤ –5D) adults aged 40-80 years, with prevalence reaching over 50% for myopia above –8D.^23^ Elsewhere, in a population-based cohort of rural Indians, PM was present in over 40% of 35 highly myopic (axial length ≥ 26mm) eyes.^24^

Older age and increasing myopia severity are two established PM risk factors.^8^ The deleterious impact of the age-related, progressive nature of PM is highlighted in recent work focusing on very old participants aged ≥ 85 years, where severe myopic maculopathy (≥ M3) was observed in almost all eyes with high myopia (axial length ≥ 26.5mm).^25^ Notably, around 60% and 90% of participants with severe myopic maculopathy exhibited at least a moderate level of binocular visual impairment (< 6/18 Snellen acuity).^25^ These findings suggest an elevated risk of visual impairment for participants in our study in later life—despite most disease presentations being mild at present with seemingly little visual impact on average (median VA ≤ 0.06 logMAR). The association between female sex and higher odds of PM, on the contrary, has not been consistently reported, with some studies^24, 26 27^ finding evidence of an association but not others.^22, 23^ A large-scale, pooled analysis of population-based studies from across Asia (with covariate adjustment) provides strong cross-sectional evidence,^28^ although longitudinal evidence remains weak.^8^ This inconclusiveness notwithstanding, the general observation that females have a higher prevalence of PM fits with the well-known male-female health-survival paradox in epidemiology, where women tend to have more morbidities despite having a longer life expectancy on average.^29^

Consistent with previous studies,^22, 26^ we found no association between education level (often used as a proxy measure for socioeconomic status) and PM, although the pooled analysis mentioned above found evidence of an association between lower education and increased odds of PM.^28^ Using a more direct measure of socioeconomic status (Townsend deprivation index), we found that lower deprivation was independently associated with higher odds of PM, which runs counter to the general expectation that deprivation leads to higher risks of morbidities due to lower exposure to health-promoting lifestyle and behavioural factors. The significance and reason for this observation remain nebulous, but it may suggest that the “environment” has a relatively limited influence on PM risk. This interpretation appears aligned with our findings that smoking, alcohol consumption, BMI and other modifiable health indicators had no influence on PM odds. Similarly, reasons for the significant difference in PM prevalence/odds between White and non-White ethnicities, even after controlling for potential confounders such as myopia severity, remain unclear. In the multi-ethnic SEED cohort, no ethnic difference was noted between Indians and Chinese/Malays after covariate adjustment. Caution should be exercised when interpretating our finding of an effect of ethnicity on PM, as unlike the SEED cohort—which had an equally large sample size for each ethnic group—the UK Biobank was not designed as a multi-ethnic cohort, thus limiting its generalisability to ethnic minorities. Given the high prevalence of PM among highly myopic Chinese adults reported by studies discussed in the preceding paragraphs, the inordinately low prevalence of PM among Chinese high myopes in the UK Biobank may reflect a high level of participation bias for ethnic minorities (perhaps healthier-than-average ethnic minorities were more likely to participate).

Few studies have investigated imaging biomarkers in PM. Previously noted OD changes include disc enlargement, tilted disc (assessed qualitatively) and a less vertical disc orientation.^21, 22^ However, it is unclear if these changes are *independent* of myopia severity^15^ due to the lack of adjustment for refractive error or axial length. For instance, in one study, an initially significant association between OD area and PM disappeared after controlling for myopia severity.^24^ Similarly, in our study, a significant association between tilted disc and PM (odds ratio: 3.49, p=0.03) was observed in the absence of covariate adjustment, but this association disappeared after controlling for covariates, including SER (Supplementary Table 1). Eyes with myopic retinopathy have previously been found to exhibit narrower central retinal arterioles and venules than myopic normal controls—even after controlling for refractive error—with arteriolar narrowing (–23.8 μm) appearing more pronounced than venular narrowing (–13.7 μm) on average.^28^ This aligns with our finding of a negative association between AVR and PM odds. Mao et al.^29^ recently reported a decrease in vessel FD from 1.45 to 1.30 as myopic maculopathy increased from M0 to M4—without adjusting for myopia severity. Thus, this does not demonstrate whether FD has independent predictive value and likely reflects FD changes due to increasing myopia from M0 to M1, as FD decreases with increasing myopia.^15^ Indeed, in our study, the mean FD for M0 was also higher (1.46) than for M3 (1.41) or M4 (1.35), reflecting increasing myopia severity from M0 to M4. After controlling for SER and other covariates in the final multivariable model, we found that PM was associated with higher, rather than lower, FD.

Unlike other region-specific population-based studies,^19–22, 24^ our analysis stands out for its country-wide scope (except Northern Ireland) through the use of the UK Biobank database. Other strengths include a significantly larger sample of high myopes (1994 analysed) compared to similar studies (≤ 626 high myopes),^19–24^ as well as the investigation of fundus biomarkers with covariate adjustment. Despite its extensive geographical coverage (depth) and the vast amount of data collected (breadth), the UK Biobank, as a volunteer-based cohort with a relatively low response rate (5.5%),^30^ is not fully representative of UK’s mid-life population. Respondents were more likely to be older, female and socioeconomically advantaged than nonparticipants.^30^ This implies that the estimated prevalence of PM is skewed towards individuals with these characteristics, likely resulting in some overestimation. Nevertheless, the database’s unique depth and breadth still provide a valuable, albeit imperfect, “first approximation” of the scale of PM in the country.

## Conclusions

To our knowledge, this is the first epidemiological study of PM in the UK, with an estimated prevalence of 41.7% among middle-aged, highly myopic participants in the UK Biobank. The lack of association between PM and any of the explored modifiable variables—except for myopia severity—underscores the need to prioritise myopia prevention and control (rather than just vision correction) as the standard of care for children or adolescents, during which eye growth is modifiable.^31, 32^ The significant associations of OD orientation, vessel FD and AVR with PM—independent of myopia severity and age—suggest that these features may be valuable adjuncts to established PM risk factors for facilitating more personalised and early prediction of PM, particularly since the associations appeared stronger at lower levels of high myopia. Future longitudinal studies should explore these associations further, as fundus features may collectively reflect the extent to which the posterior pole is stretched in individual eyes.

## Data Availability

Data directly supporting the results of this work are only available to the immediate research team members due to UK Biobank's access control policy. Bona fide researchers can, however, apply for access at ukbiobank.ac.uk/enable-your-research/apply-for-access. Source code used to perform the analysis described in this work is freely available at github.com/fyii200/UKbiobankPM.

https://ukbiobank.ac.uk/enable-your-research/apply-for-access

## Acknowledgements

The authors would like to thank the Belfast Ophthalmic Reading Centre for grading the fundus photographs analysed in this study. F. Yii acknowledges funding support from: the Medical Research Council (MR/N013166/1 & Flexible Supplement Funds), the Edinburgh Futures Institute, and the Visual Research Trust (SCOO4516). M.O. Bernabeu acknowledges funding support from: Fondation Leducq Transatlantic Network of Excellence (17 CVD 03); EPSRC grant no. EP/X025705/1; British Heart Foundation and The Alan Turing Institute Cardiovascular Data Science Award (C-10180357); Diabetes UK (20/0006221); Fight for Sight (5137/5138); the SCONe projects funded by Chief Scientist Office, Edinburgh & Lothians Health Foundation, Sight Scotland, the Royal College of Surgeons of Edinburgh, the RS Macdonald Charitable Trust, and Fight For Sight.

## References

1. Haarman AEG, Enthoven CA, Tideman JWL, et al. The complications of myopia: A review and meta-analysis. Invest Ophthalmol Vis Sci 2020;61(4):49. doi: 10.1167/iovs.61.4.49

2. Ohno-Matsui K, Wu PC, Yamashiro K, et al. IMI Pathologic Myopia. Invest Ophthalmol Vis Sci 2021;62(5):5. doi: 10.1167/iovs.62.5.5

3. Duke-Elder S, Abrams D. System of Ophthalmology: Ophthalmic Optics and Refraction. London: Kimpton 1970:300–301.

4. Sorsby A. Blindness in England 1951-1954. London: H.M. Stationery Office 1950.

5. Naidoo KS, Fricke TR, Frick KD, et al. Potential lost productivity resulting from the global burden of myopia: Systematic review, meta-analysis, and modeling. Ophthalmology 2019;126(3):338–46. doi: 10.1016/j.ophtha.2018.10.029 [published Online First: 20181017]

6. Wong YL, Saw SM. Epidemiology of Pathologic Myopia in Asia and Worldwide. Asia Pac J Ophthalmol (Phila) 2016;5(6):394–402. doi: 10.1097/APO.0000000000000234

7. Zou M, Wang S, Chen A, et al. Prevalence of myopic macular degeneration worldwide: A systematic review and meta-analysis. Br J Ophthalmol 2020;104(12):1748–54. doi: 10.1136/bjophthalmol-2019-315298 [published Online First: 20200318]

8. Yii F, Nguyen L, Strang N, et al. Factors associated with pathologic myopia onset and progression: A systematic review and meta-analysis. Ophthalmic Physiol Opt 2024 doi: 10.1111/opo.13312 [published Online First: 20240402]

9. Chua SYL, Thomas D, Allen N, et al. Cohort profile: design and methods in the eye and vision consortium of UK Biobank. BMJ Open 2019;9(2):e025077. doi: 10.1136/bmjopen-2018-025077 [published Online First: 20190221]

10. Shen D, Liu T, Peters TM, et al., eds. Evaluation of Retinal Image Quality Assessment Networks in Different Color-Spaces. Medical Image Computing and Computer Assisted Intervention – MICCAI 2019; 2019 2019//; Cham. Springer International Publishing.

11. Ohno-Matsui K, Kawasaki R, Jonas JB, et al. International photographic classification and grading system for myopic maculopathy. Am J Ophthalmol 2015;159(5):877–83.e7. doi: 10.1016/j.ajo.2015.01.022 [published Online First: 20150126]

12. Yokoi T, Jonas JB, Shimada N, et al. Peripapillary Diffuse Chorioretinal Atrophy in Children as a Sign of Eventual Pathologic Myopia in Adults. Ophthalmology 2016;123(8):1783–87. doi: 10.1016/j.ophtha.2016.04.029 [published Online First: 20160522]

13. Townsend P, Phillimore P, Beattie A. Health and Deprivation: Inequality and the North. London: Routledge 1988.

14. UK Biobank. First Occurrence of Health Outcomes Defined by 3-character ICD10 code. 2019. https://biobank.ndph.ox.ac.uk/showcase/ukb/docs/first_occurrences_outcomes.pdf.

15. Yii F, Bernabeu MO, Dhillon B, et al. Retinal Changes From Hyperopia to Myopia: Not All Diopters Are Created Equal. Invest Ophthalmol Vis Sci 2024;65(5):25. doi: 10.1167/iovs.65.5.25

16. Yii F, Strang N, Moulson C, et al. The optical nature of myopic changes in retinal vessel calibre. Ophthalmology Science 2024:100631. doi: 10.1016/j.xops.2024.100631

17. Jonas JB, Kling F, Gründler AE. Optic disc shape, corneal astigmatism, and amblyopia. Ophthalmology 1997;104(11):1934–7. doi: 10.1016/s0161-6420(97)30004-9

18. Thompson CG, Kim RS, Aloe AM, et al. Extracting the Variance Inflation Factor and Other Multicollinearity Diagnostics from Typical Regression Results. Basic and Applied Social Psychology 2017;39(2):81–90. doi: 10.1080/01973533.2016.1277529

19. Haarman AEG, Tedja MS, Brussee C, et al. Prevalence of Myopic Macular Features in Dutch Individuals of European Ancestry With High Myopia. JAMA Ophthalmol 2022;140(2):115–23. doi: 10.1001/jamaophthalmol.2021.5346

20. Hopf S, Korb C, Nickels S, et al. Prevalence of myopic maculopathy in the German population: results from the Gutenberg health study. Br J Ophthalmol 2020;104(9):1254–59. doi: 10.1136/bjophthalmol-2019-315255 [published Online First: 20191216]

21. Vongphanit J, Mitchell P, Wang JJ. Prevalence and progression of myopic retinopathy in an older population. Ophthalmology 2002;109(4):704–11. doi: 10.1016/s0161-6420(01)01024-7

22. Liu HH, Xu L, Wang YX, et al. Prevalence and progression of myopic retinopathy in Chinese adults: The Beijing Eye Study. Ophthalmology 2010;117(9):1763–8. doi: 10.1016/j.ophtha.2010.01.020 [published Online First: 20100505]

23. Wong YL, Sabanayagam C, Ding Y, et al. Prevalence, risk factors, and impact of myopic macular degeneration on visual impairment and functioning among adults in Singapore. Invest Ophthalmol Vis Sci 2018;59(11):4603–13. doi: 10.1167/iovs.18-24032

24. Jonas JB, Nangia V, Gupta R, et al. Prevalence of myopic retinopathy in rural Central India. Acta Ophthalmol 2017;95(5):e399–e404. doi: 10.1111/aos.13301 [published Online First: 20161117]

25. Bikbov MM, Gilmanshin TR, Kazakbaeva GM, et al. Prevalence of Myopic Maculopathy Among Adults in a Russian Population. JAMA Netw Open 2020;3(3):e200567. doi: 10.1001/jamanetworkopen.2020.0567 [published Online First: 20200302]

26. Chen SJ, Cheng CY, Li AF, et al. Prevalence and associated risk factors of myopic maculopathy in elderly Chinese: The Shihpai eye study. Invest Ophthalmol Vis Sci 2012;53(8):4868–73. doi: 10.1167/iovs.12-9919 [published Online First: 20120724]

27. Asakuma T, Yasuda M, Ninomiya T, et al. Prevalence and risk factors for myopic retinopathy in a Japanese population: the Hisayama Study. Ophthalmology 2012;119(9):1760–5. doi: 10.1016/j.ophtha.2012.02.034 [published Online First: 20120510]

28. Wong YL, Zhu X, Tham YC, et al. Prevalence and predictors of myopic macular degeneration among Asian adults: Pooled analysis from the Asian Eye Epidemiology Consortium. Br J Ophthalmol 2021;105(8):1140–48. doi: 10.1136/bjophthalmol-2020-316648 [published Online First: 20200902]

29. Oksuzyan A, Petersen I, Stovring H, et al. The male-female health-survival paradox: a survey and register study of the impact of sex-specific selection and information bias. Ann Epidemiol 2009;19(7):504–11. doi: 10.1016/j.annepidem.2009.03.014 [published Online First: 20090519]

30. Fry A, Littlejohns TJ, Sudlow C, et al. Comparison of Sociodemographic and Health-Related Characteristics of UK Biobank Participants With Those of the General Population. Am J Epidemiol 2017;186(9):1026–34. doi: 10.1093/aje/kwx246

31. Yii FS. Emmetropic eye growth in East Asians and non-East Asians. Ophthalmic Physiol Opt 2023;43(6):1412–18. doi: 10.1111/opo.13195 [published Online First: 20230627]

32. Naduvilath T, He X, Saunders K, et al. Regional/ethnic differences in ocular axial elongation and refractive error progression in myopic and non-myopic children. Ophthalmic Physiol Opt 2024 doi: 10.1111/opo.13401 [published Online First: 20241007]

